# Feasibility Study on the Application of Transcranial Focused Ultrasound Stimulation in Ischemic Stroke Rehabilitation

**DOI:** 10.1101/2024.11.13.24317114

**Authors:** JeYoung Jung, Minjae Cho, Sekwang Lee, Hyun-Joon Yoo, Matthew A. Lambon Ralph, Sung-Bom Pyun

## Abstract

Stroke is a leading cause of long-term neurological disability and death worldwide. Traditional neurorehabilitation methods result in full recovery for less than 15% of stroke patients, highlighting the need for innovative approaches. Low-intensity transcranial focused ultrasound stimulation (tFUS) has emerged as a promising non-invasive neuromodulation technique with high focality and deep tissue penetration. This study evaluates the feasibility, safety and efficacy of tFUS for motor rehabilitation in ischemic stroke patients. We conducted a single-group prospective pilot study involving two stroke patients who received tFUS targeting the motor cortex contralateral to the lesion site three times a week. Patients were evaluated using multiple clinical measures of motor function. Both patients showed improvements in the Fugl-Meyer Assessment upper extremity score, kinesthetic sensation, stereognosis score from the Nottingham Sensory Assessment, and the Modified Barthel Index score. Follow-up evaluations immediately and three weeks after stimulation indicated sustained motor function improvements. The results suggest that tFUS can effectively enhance motor recovery in stroke patients without adverse events. This study provides robust evidence supporting the potential of tFUS in stroke rehabilitation, opening new avenues for clinical application and research.

## Introduction

Stroke is a leading cause of long-term neurological disability and the second leading cause of death globally, affecting over 13.7 million people and causing 5.7 million deaths annually ^1^. Despite traditional neurorehabilitation approaches, less than 15% of stroke patients fully recover, highlighting the need for innovative neurotechnology-based strategies to enhance recovery ^2^. Most strokes are ischemic, often affecting the middle cerebral artery (MCA) and resulting in damage to critical brain regions such as the primary somatosensory and motor cortices, basal ganglia, thalamus, caudate, and internal capsule ^3^. Given that most stroke lesions include subcortical, there is a clear need for neuromodulation techniques capable of reaching deep brain regions for effective post-stroke rehabilitation.

Low-intensity transcranial focused ultrasound stimulation (tFUS) has emerged as a promising non-invasive technique for neuromodulation, capable of safely and painlessly targeting deep brain regions with high focality and penetrability ^4^. Pre-clinical studies in animals and initial human studies suggest potential safety and feasibility of tFUS with minimal adverse effects ^5^. tFUS can modulate neural excitability through mechanical effects, such as changes in ion channels ^6^, alterations in membrane capacitance ^7^, and temperature increases caused by the ultrasound waves ^8^. Recently, tFUS has shown its applicability in treating neurological and psychiatric disorders, including Parkinson’s disease ^9^, Alzheimer’s disease ^10^, epilepsy ^11^ and depression ^12^ in humans.

Despite its potential, there are no human studies on tFUS for stroke rehabilitation. However, animal studies show promising results. For example, Wu et al. ^13^ demonstrated that tFUS targeted at the cortical penumbra improved outcomes in rats with endothelin-1 induced MCA occlusion strokes. Kim et al. ^14^ developed a wearable tFUS system for rats with MCA occlusion strokes, targeting cortical and subcortical regions and showing enhanced rehabilitation outcomes. Another study ^15^ showed that tFUS enhanced neurological repair and remodeling in ischemic stroke by increasing cerebral blood flow and promoting angiogenesis in mice.

In this study, we evaluated the feasibility of tFUS neuromodulation in stroke rehabilitation. This clinical pilot study focused on patients with ischemic stroke to assess the feasibility, tolerability and efficacy of tFUS in promoting motor recovery. tFUS was applied to the motor cortex contralateral to the lesion site to modulate the motor system. Building on promising animal studies, we investigated the potential of tFUS to improve motor function in these patients.

## Materials and Methods

### Participants

This study was a single-group prospective pilot study with two stroke patients. The patients were selected by following inclusion criteria; 1) adults aged 19 to 85 years, 2) patients diagnosed with their first stroke confirmed by brain MRI, 3) patients in the subacute phase within 3 months of an ischemic stroke occurrence at the time of screening, 4) patients with persistent unilateral motor paralysis due to a stroke at the time of screening: specifically, patients with a unilateral upper limb muscle strength grade of 4 (good grade) or lower, as evaluated by Medical Research Council manual muscle testing (MMT), 5) patients who were conscious, able to cooperate with the study, and provided voluntary consent to participate in this clinical trial.

Additionally, the patients were excluded by the following criteria: 1) women who were pregnant or breastfeeding (for women of childbearing potential, a positive urine HCG test); 2) patients with any of the following conditions observed on brain imaging (CT or MRI): skull thickness of 8 mm or more, meningioma, brain tumor, abscess, fluid retention, hydrocephalus, trauma, skull fracture, congenital brain malformation, or any other cerebral hemisphere abnormalities or history of brain diseases other than ischemic stroke; 3) patients with abnormal results in the basic hematological screening tests including renal disease (creatinine > 2.0 mg/dl), thrombocytopenia (platelets < 100 × 10^3^/μl), leukopenia (white blood cells < 4.0 × 10^3^/μl), liver dysfunction (AST, ALT, total bilirubin levels more than twice the upper limit of the normal range).

This study was approved by the Institutional Review Board of Korea University Anam Hospital (approval no. 2023AN0046) and was registered with the Clinical Research Information Service (registration no. KCT0008320).

The details of demographic and clinical characteristics of the two participants are summarized in Table 1 and the MRI images of the two patients are shown in Figure 1. Korean version of the Mini-Mental State Examination (K-MMSE) scores indicated that Patient 1 had cognitive impairment, with a score of 21 out of a possible 30 points. Patient 2, however, showed normal cognitive function. The Hospital Anxiety and Depression Scale (HADS) scores indicate that scores of 8 or above for both anxiety and depression suggest the presence of symptoms. Both patients had depression scores within the normal range and mild anxiety.

**Table 1.** Demographic characteristics of the participants.

| Characteristics | Patient 1 | Patient 2 |
| --- | --- | --- |
| Age range | 71-75 | 65-70 |
| Sex | Female | Male |
| Lesion | Right. MCA | Left. BG & CR |
| Time after onset (days) | 47 | 41 |
| Education(years) | 6 | 12 |
| Handedness | Right | Right |
| K-MMSE | 21 | 28 |
| HADS |  |  |
| HAD-A | 8 | 10 |
| HAD-B | 4 | 6 |
| MMT |  |  |
| Upper | 4~4+ | 0~3 |
|  | 1~2- | 5 |
| Lower | 4+ | 5- |
|  | 2+~3- | 5 |

Abbreviations: MCA, Middle cerebral artery; BG, Basal ganglia; CR, Corona radiata; K-MMSE, Korean version of the Mini-Mental State Examination; HADS, Hospital Anxiety and Depression Scale (HADS); MMT, Manual Muscle Testing

**Figure 1.**
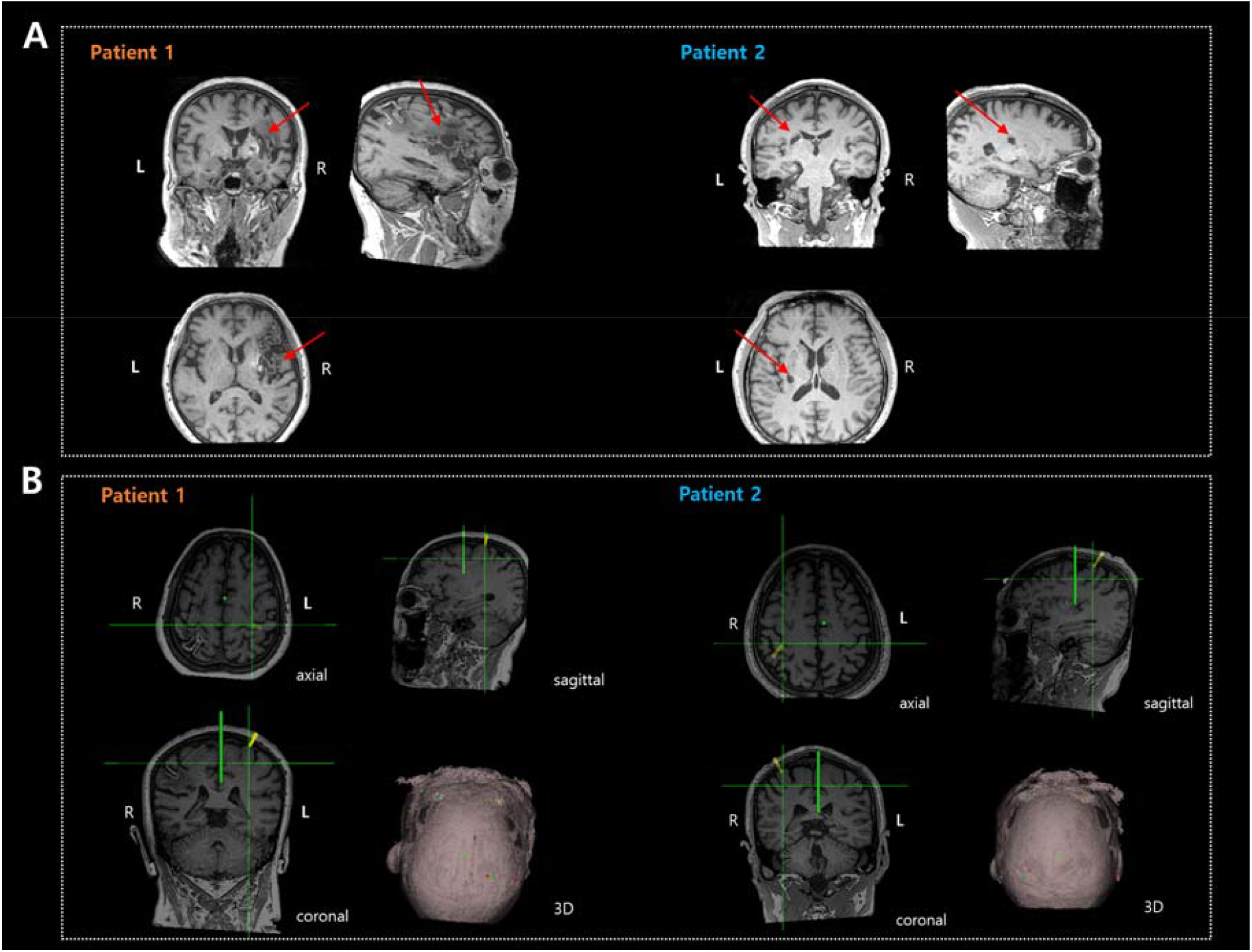
A) Structural images of the two patients. Red arrows indicate lesion sites. B) MRI images of entry and target points (yellow line) for the two stroke patients.

### Therapeutic Focused Ultrasound device

This therapeutic focused ultrasound stimulation (FUS) system, NS-US200 device (Neurosona Co., Ltd., Seoul, Korea), uses low-intensity FUS (ranges from 10 mW/cm^2^ to 50 W/cm^2^) by applying electrical signals to a focused ultrasound transducer, which then stimulates specific regions of the brain. Unlike high-intensity focused ultrasound (HIFU) systems that use thermal energy, the ultrasound stimulation used here employs mechanical energy in the form of pressure waves. It operates at a low frequency of 250 kHz and delivers low-intensity energy below 50 W/cm^2^ for brain stimulation.

The therapeutic FUS device used in this clinical trial was administered three times a week. The two selected patients participated in the FUS experiment, using the NS-US200 device with a transducer (E40) having a focal length of 30 mm and a focal size was 4 mm in diameter and 18 mm in length at 90% maximum intensity. To enhance ultrasound permeability, a hydrating gel was applied to the patient’s scalp. Based on acquired anatomical brain imaging, the focused ultrasound was non-invasively administered to the central sulcus ‘knob’ area within the primary motor cortex (M1) contralateral to the lesion site using Neurosona Ultrasound Software (Fig. 1B).

We set the low-intensity focused ultrasound parameters with a repetition frequency (PRF) of 100 Hz and an acoustic intensity at focus (AIF) of 3 W/cm^2^. The Tone Burst Duration (TBD) of the stimulation parameters was 0.5 msec, with a duty cycle of 5%. The total treatment duration was 1200 seconds without a sonication interval. The *in situ* mechanical index (MI) was maintained at 0.6, the fundamental frequency was 250 kHz, and the spatial peak temporal-average acoustic intensity (I_SPTA_) was 1.5 W/cm^2^.

Using the open-source software application 3D Slicer (https://www.slicer.org/), we performed preprocessing and registration of T1-weighted MRI images and brain CT images. Before the MRI scans, each patient had four fiducial markers attached to the following regions: the left and right brow bones, and the mastoid bones located behind the ears. For validation, we used NUS viewer (Neurosona software) to set the four fiducial marker coordinates for each patient before brain stimulation. We used the registered images to target the intact hemisphere’s primary motor cortex (M1) at the hand knob, which is shaped like an omega. For patient 1, who had a lesion in the right hemisphere, stimulation was applied to the intact left hemisphere. Conversely, for patient 2, who had a lesion in the left hemisphere, stimulation was applied to the right hemisphere (Fig. 1B).

### Clinical Assessment

Before brain stimulation, we collected clinical information about participant’s age, gender, location of stroke, date of onset, dominant hand and neurological assessment.

The procedures for the entire clinical trials are summarized in Table 2. For all assessments, higher scores indicated better performance. In the baseline assessments, the participants completed the K-MMSE, Hospital Anxiety scale and HADS. The HADS is a 14 item self-report scale with each 7 item subscales for anxiety and depression with total score from 0–42. Additionally, we included clinical assessments for motor function. The MMT measures extremity muscle strength which grades muscle strength from 0 to 5, with grade 5 indicating normal strength. The Fugl-Meyer Assessment (FMA) evaluates balance, sensation and joint of motion for those who have had a stroke. The FMA consists of upper and lower extremity scores, 66 points and 34 points respectively, with higher scores indicating better performance. The Finger Grip Strength test uses a dynamometer to measure grip force, conducted three times, with the score being the average of the three attempts. The Manual Function Test (MFT) assesses upper limb function that maximum scores are 32 points. The Action Research Arm Test (ARAT) is used to measure upper extremity function and consists of 19 items, with total scores ranging from 0 to 57; higher scores indicate better performance. The Box and Block Test assesses unilateral gross manual dexterity by requiring individuals to move miniature blocks from one compartment of a box to another as many times as possible within 60-seconds. The Berg Balance Scale (BBS) is a test for balance function and the total score is 56 with higher scores representing better balance function. Nottingham Sensory Assessment is an tool used to assess sensory impairment. This test assesses tactile, kinaesthetic and stereognosis sensory function. Functional Ambulation Categories (FAC) assesses ambulation ability by measuring how well a patient can walk without assistance. The Modified Barthel Index (MBI) evaluates dependence in activities of daily living and consists of categories of basic care tasks, such as feeding, bathing, dressing, grooming, toileting, transfer, ambulation and stair climbing.

**Table 2.** Summary of clinical assessments before and after tFUS.

| Assessment | Patient 1 |  |  | Patient 2 |  |  |
| --- | --- | --- | --- | --- | --- | --- |
|  | Baseline | Post1<br>(5days) | Post2<br>(22 days) | Baseline | Post1<br>(5days) | Post2<br>(22days) |
| FMA |  |  |  |  |  |  |
| Upper | 6 | 7 | 8 | 12 | 14 | 15 |
| Lower | 13 | 13 | 13 | 34 | 34 | 34 |
| BBS | 4 | 4 | 4 | 52 | 52 | 54 |
| Nottingham Sensory |  |  |  |  |  |  |
| Tactile sensation | 15~18 | 15~18 | 14~18 | 18 | 18 | 18 |
| Kinaesthetic sensation | 17 | 19 | 19 | 21 | 21 | 21 |
| Stereognosis | 3 | 4 | 4 | 22 | 22 | 22 |
| FAC | 0 | 0 | 0 | 5 | 5 | 5 |
| MBI | 43 | 43 | 52 | 88 | 88 | 91 |
| Finger Grip |  |  |  |  |  |  |
| Left | NT | NT | NT | 34.8 | 37.3 | 35.9 |
| Right | 24.4 | 20.3 | 19.4 | NT | NT | NT |
| ARAT |  |  |  |  |  |  |
| Left | 0 | 0 | 0 | 57 | 57 | 57 |
| Right | 56 | 57 | 57 | 4 | 4 | 4 |
| Box and Block |  |  |  |  |  |  |
| Non-paralyzed side | 42 | 53 | 50 | 63 | 67 | 69 |

Abbreviations: FMA, Fugl-Meyer Assessment; BBS, Berg Balance Scale; FAC, Functional Ambulation Categories; MBI, Modified Barthel Index; ARAT, Action Research Arm Test

The clinical assessments were conducted at 3-time points; pre-stimulation (Baseline) and immediately after completion of FUS (Post1) and 3 weeks after the initial brain stimulation (Post2).

Adverse event evaluations, including stroke exacerbation, seizure occurrence, headaches, and other neurological symptoms, were conducted at each stimulation session as well as at 3 weeks.

## Results

There was no adverse event in either participant during or after the brain stimulations. Both participants showed an improvement in the FMA upper extremity score, kinaesthetic sensation and stereognosis score of Nottingham Sensory Assessment and MBI score. Additionally, Patient 2 demonstrated improved scores on the BBS and Box and Block tests. The results are summarized in Table 2.

For follow-up evaluations, both patients showed slight improvements in the FMA upper extremity scores. However, both scored below 30 out of a possible 66 points, indicating severe impairment (mild: <45, moderate: 30-45, severe: <30). In the lower extremity evaluation, Patient 2 maintained a normal status with perfect scores out of 34 points, while Patient 1 continued to show severe disability, scoring the same as at baseline.

The BBS results indicated that both patients either maintained their baseline status or, in the case of Patient 2, showed slight improvement following brain stimulation. Patient 1 remained at a greater risk of falling due to lower extremity paralysis, whereas Patient 2 exhibited a significantly lower risk of falling.

The Nottingham Sensory Assessment and FAC evaluation results showed no notable changes compared to the baseline results. MBI evaluations showed that both patients, initially categorized as severe and moderate, respectively, demonstrated gradual improvement over time. The finger grip assessment was conducted on the intact side only, and the results were within the normal range according to gender-specific norms. In the ARAT, both the baseline and follow-up evaluations showed that the intact side scored a perfect 57 points, indicating high ability (Grattan, Emily S et al. 2019). In the Box and Block Test, Patient 2 showed near-normal performance, while Patient 1 showed slightly lower performance. However, both patients demonstrated improved performance in the follow-up evaluation compared to baseline.

## Discussion

tFUS holds significant promise for treating ischemic stroke due to its safety, non-invasiveness, deep tissue penetration, and high spatial precision ^16^. In this study, we assessed the feasibility of using tFUS for motor recovery rehabilitation in patients with ischemic stroke. We demonstrated feasibility through successful targeting and the ability of all patients to complete the treatment without adverse events. The outcome data revealed efficacy in several clinical measurements of motor function, reinforcing the safety of tFUS in ischemic stroke. Importantly, our results indicated both immediate and long-term benefits of tFUS for stroke motor recovery. This study provides robust evidence supporting the feasibility, safety, and efficacy of tFUS in stroke rehabilitation.

Various non-invasive brain stimulation (NIBS) techniques have been used to promote functional recovery after stroke, with proven efficacy across various functions, including motor, cognitive, language and swallowing abilities ^17-20^. In this study, we adjusted the ultrasound parameters to a PRF of 100Hz with a duty cycle of 5% for 20 minutes to attempt contralesional inhibitory stimulation ^21^. The approach of stimulating and inhibiting the contralesional hemisphere has been widely applied in other NIBS techniques, and its conceptual foundation can be explained by the principle of “interhemispheric inhibition” ^22,23^. In a healthy brain, there is a reciprocal inhibitory interaction between the hemispheres through pathways crossing the corpus callosum, especially in the motor system. When a stroke occurs, the inhibitory function of the affected hemisphere is reduced, leading to an increase in inhibitory activity in the contralesional hemisphere ^24,25^. Therefore, in this study, we stimulated the contralesional primary motor cortex (M1 cortex) through inhibitory ultrasound stimulation to enhance the functional activation of the affected hemisphere.

Although we attempted inhibitory ultrasound stimulation, the precise mechanisms underlying the excitatory and inhibitory effects of tFUS remain unclear. Several mechanisms may contribute to the neuromodulatory effects of tFUS such as membrane displacement linked to capacitance changes, the sonoporation effect, and activation of mechanosensitive channels ^8^. Different studies have reported various effects, leading to the proposal of models such as the neuronal bilayer sonophore model ^7^ and the neuronal intramembrane cavitation excitation model ^26^. These models suggest that ultrasound can selectively activate different cortical neuron subtypes, including excitatory regular spiking pyramidal neurons, inhibitory fast-spiking cortical neurons, and inhibitory low-threshold spiking cortical neurons. The overall neuromodulation effects (excitation or suppression) are determined by the interactions between these selectively activated excitatory and inhibitory neurons. Recent evidence from Yu et al.^27^ indicates that by adjusting the pulse PRF of the ultrasound, specific neuron types can be preferentially targeted in vivo in anesthetized rodent brains. However, the precise mechanisms of tFUS modulation on the M1 and its network effects in stroke require further study.

Despite these uncertainties and the conceptual similarities with other NIBS techniques, confirming tFUS’s feasibility, efficacy and safety in stroke patients in this study suggests that tFUS may offer distinct advantages over other NIBS methods. For instance, tFUS has the benefits of deeper penetration and higher spatial precision, allowing it to reach deeper brain structures that were previously inaccessible while minimizing the impact on surrounding tissues ^8,28^. This capability raises the potential for personalized treatment, allowing the intensity and location of the stimulation to be tailored to the patient’s specific symptoms, thereby providing a more targeted and effective intervention to improve post-stroke impairment.

Nonetheless, it is crucial to recognize that, as a pilot study, we cannot conclusively determine whether the observed improvements in motor function are directly due to tFUS. This limitation highlights the necessity for additional research with larger sample sizes and appropriate control groups to more precisely assess the direct impact of tFUS on motor recovery following a stroke.

Given these findings, the potential of tFUS in rehabilitation opens new avenues for research and therapeutic applications. Future studies should focus on optimizing stimulation parameters, identifying the most effective brain regions for different cognitive functions, and exploring long-term effects and safety. Additionally, combining tFUS with other cognitive training or therapeutic interventions could synergistically enhance its efficacy.

## Data Availability

All data produced in the present study are available upon reasonable request to the authors.

## Data availability

Data will be available from the corresponding author upon reasonable request.

## Acknowledgements

We acknowledge two patients who participated in this study.

## Author Contributions

Pyun S.B and Yoo H.J. designed the experiment; Lee S.K., and Cho M.J. performed the study; Cho M.J. and Jung J. analyzed the results; Cho M.J. Jung J, Lee S.K, Lambon Ralph M.A., and Pyun S.B. wrote the paper. All authors have read and approved the final version of the manuscript.

## References

1. eclinicalmedicine. The rising global burden of stroke. Eclinicalmedicine. May 2023;59 doi:ARTN 102028 10.1016/j.eclinm.2023.102028

2. Grefkes C, Fink GR. Recovery from stroke: current concepts and future perspectives. Neurol Res Pract. Jun 16 2020;2(1) doi:ARTN 17 10.1186/s42466-020-00060-6

3. Nichols L, Bridgewater JC, Wagner NB, et al. Where in the Brain do Strokes Occur? A Pilot Study and Call for Data. Clin Med Res. Sep 1 2021;19(3):110–115. doi:10.3121/cmr.2021.1632

4. Darmani G, Bergmann TO, Pauly KB, et al. Non-invasive transcranial ultrasound stimulation for neuromodulation. Clin Neurophysiol. Mar 2022;135:51–73. doi:10.1016/j.clinph.2021.12.010

5. Pasquinelli C, Hanson LG, Siebner HR, Lee HJ, Thielscher A. Safety of transcranial focused ultrasound stimulation: A systematic review of the state of knowledge from both human and animal studies. Brain Stimul. Nov-Dec 2019;12(6):1367–1380. doi:10.1016/j.brs.2019.07.024

6. Yoo S, Mittelstein DR, Hurt R, Lacroix J, Shapiro MG. Focused ultrasound excites cortical neurons via mechanosensitive calcium accumulation and ion channel amplification. Nat Commun. Jan 25 2022;13(1) doi:ARTN 493 10.1038/s41467-022-28040-1

7. Plaksin M, Shoham S, Kimmel E. Intramembrane cavitation as a predictive bio-piezoelectric mechanism for ultrasonic brain stimulation. J Mol Neurosci. Aug 2014;53:S103–S103.

8. Blackmore J, Shrivastava S, Sallet J, Butler CR, Cleveland RO. Ultrasound Neuromodulation: A Review of Results, Mechanisms and Safety. Ultrasound Med Biol. Jul 2019;45(7):1509–1536. doi:10.1016/j.ultrasmedbio.2018.12.015

9. Osou S, Radjenovic S, Bender L, et al. Novel ultrasound neuromodulation therapy with transcranial pulse stimulation (TPS) in Parkinson’s disease: a first retrospective analysis. J Neurol. Mar 2024;271(3):1439–1450. doi:10.1007/s00415-023-12114-1

10. Beisteiner R, Matt E, Fan C, et al. Transcranial Pulse Stimulation with Ultrasound in Alzheimer’s Disease-A New Navigated Focal Brain Therapy. Adv Sci. Feb 2020;7(3) doi:ARTN 1902583 10.1002/advs.201902583

11. Stern JM, Spivak NM, Becerra SA, et al. Safety of focused ultrasound neuromodulation in humans with temporal lobe epilepsy. Brain Stimul. Jul-Aug 2021;14(4):1022–1031. doi:10.1016/j.brs.2021.06.003

12. Riis TS, Feldman DA, Vonesh LC, et al. Durable effects of deep brain ultrasonic neuromodulation on major depression: a case report. J Med Case Rep. Oct 28 2023;17(1) doi:ARTN 449 10.1186/s13256-023-04194-4

13. Wu CT, Yang TH, Chen MC, et al. Low Intensity Pulsed Ultrasound Prevents Recurrent Ischemic Stroke in a Cerebral Ischemia/Reperfusion Injury Mouse Model via Brain-derived Neurotrophic Factor Induction. Int J Mol Sci. Oct 2 2019;20(20) doi:ARTN 5169 10.3390/ijms20205169

14. Kim E, Anguluan E, Kum J, et al. Wearable Transcranial Ultrasound System for Remote Stimulation of Freely Moving Animal. Ieee T Bio-Med Eng. Jul 2021;68(7):2195–2202. doi:10.1109/Tbme.2020.3038018

15. Qi L, Wang C, Deng LD, et al. Low-intensity focused ultrasound stimulation promotes stroke recovery via astrocytic HMGB1 and CAMK2N1 in mice. Stroke Vasc Neurol. Jan 8 2024; doi:10.1136/svn-2023-002614

16. Guo JC, Lo WLA, Hu HJ, Yan L, Li L. Transcranial ultrasound stimulation applied in ischemic stroke rehabilitation: A review. Front Neurosci-Switz. Jul 22 2022;16 doi:ARTN 964060 10.3389/fnins.2022.964060

17. Wessel MJ, Zimerman M, Hummel FC. Non-invasive brain stimulation: an interventional tool for enhancing behavioral training after stroke. Front Hum Neurosci. 2015;9:265. doi:10.3389/fnhum.2015.00265

18. Draaisma LR, Wessel MJ, Hummel FC. Non-invasive brain stimulation to enhance cognitive rehabilitation after stroke. Neurosci Lett. Feb 6 2020;719:133678. doi:10.1016/j.neulet.2018.06.047

19. Thiel A, Hartmann A, Rubi-Fessen I, et al. Effects of noninvasive brain stimulation on language networks and recovery in early poststroke aphasia. Stroke. Aug 2013;44(8):2240–6. doi:10.1161/STROKEAHA.111.000574

20. Cheng I, Sasegbon A, Hamdy S. Effects of Neurostimulation on Poststroke Dysphagia: A Synthesis of Current Evidence From Randomized Controlled Trials. Neuromodulation. Dec 2021;24(8):1388–1401. doi:10.1111/ner.13327

21. Dell’Italia J, Sanguinetti JL, Monti MM, Bystritsky A, Reggente N. Current State of Potential Mechanisms Supporting Low Intensity Focused Ultrasound for Neuromodulation. Frontiers in Human Neuroscience. Apr 25 2022;16 doi:ARTN 872639 10.3389/fnhum.2022.872639

22. Gorsler A, Bäumer T, Weiller C, Münchau A, Liepert J. Interhemispheric effects of high and low frequency rTMS in healthy humans. Clin Neurophysiol. Oct 2003;114(10):1800–1807. doi:10.1016/S1388-2457(03)00157-3

23. Pal PK, Hanajima R, Gunraj CA, et al. Effect of low-frequency repetitive transcranial magnetic stimulation on interhemispheric inhibition. J Neurophysiol. Sep 2005;94(3):1668–1675. doi:10.1152/jn.01306.2004

24. Murase N, Duque J, Mazzocchio R, Cohen LG. Influence of interhemispheric interactions on motor function in chronic stroke. Ann Neurol. Mar 2004;55(3):400–409. doi:10.1002/ana.10848

25. Alia C, Spalletti C, Lai S, et al. Neuroplastic Changes Following Brain Ischemia and their Contribution to Stroke Recovery: Novel Approaches in Neurorehabilitation. Front Cell Neurosci. Mar 16 2017;11 doi:ARTN 76 10.3389/fncel.2017.00076

26. Plaksin M, Kimmel E, Shoham S. Cell-Type-Selective Effects of Intramembrane Cavitation as a Unifying Theoretical Framework for Ultrasonic Neuromodulation. Eneuro. May-Jun 2016;3(3) doi:ARTN 0136-15.2016 10.1523/Eneuro.0136-15.2016

27. Yu K, Niu XD, Krook-Magnuson E, He B. Intrinsic functional neuron-type selectivity of transcranial focused ultrasound neuromodulation. Nat Commun. May 4 2021;12(1) doi:ARTN 2519 10.1038/s41467-021-22743-7

28. Fomenko A, Neudorfer C, Dallapiazza RF, Kalia SK, Lozano AM. Low-intensity ultrasound neuromodulation: An overview of mechanisms and emerging human applications. Brain Stimul. Nov-Dec 2018;11(6):1209–1217. doi:10.1016/j.brs.2018.08.013

